# Classifying Adverse Events from SOAP Notes and Sensor Features in a Clinical Trial of Older Adults

**DOI:** 10.1101/2025.08.20.25334088

**Authors:** Noah Marchal, Mihail Popescu, Erin L. Robinson, Rachel Wolpert, Xing Song

## Abstract

Early detection of adverse events and fall injuries may improve patient safety outcomes for clinical trials in geriatric populations. This study evaluates multimodal models combining structured SOAP notes and remote biophysical sensor measurements to classify adverse event occurrences and fall events in a clinical trial with rural older adults participants. XGBoost classifiers were trained on BioBERT, BioClinicalBERT and BERT-Uncased SOAP note embeddings, with and without fused sensor features, and compared across control and intervention cohorts. Non-fused embedding features performed best on Subjective notes for adverse event classification from BioClinicalBERT (AUROC=0.89, Recall=0.88) for controls and BioBERT (AUROC=0.86, Recall=0.73) in the intervention arm. Sensor features provided higher discrimination and recall for adverse events in controls (AUROC=0.68, Recall=0.80) than the intervention arm (AUROC=0.57, Recall=0.10). For fall classification, sensor features outperformed embeddings in the control (AUROC=0.87, Recall=0.32) and intervention (AUROC=0.84, Recall=0.12) cohorts. Assessment and Planning note components had significantly lower AUROC across all embedding feature models. Fusing sensor and embedding features resulted in near-perfect performance from Subjective and Objective notes (AUROC=1.0, Recall=1.0), significantly better than non-fused embeddings. Analysis of NER tokens extracted from SOAP notes showed that model performance differences are associated with cohort-specific documentation practices. SOAP contents in the intervention cohort were more patient-focused, with higher word counts in Subjective sections and narrower AUROC confidence intervals, reflecting increased clinical engagement and improved event capture. These results suggest that combining clinical narratives with continuous sensor measurements can improve the prediction of adverse events and fall injuries, which may increase clinical trial safety and reduce the frequency of in-person assessments.

## 1. Introduction

Rural older adults experience higher levels of social isolation and vulnerability, which contribute to worsening chronic conditions and disabilities due to longer intervals between healthcare visits (Pickering et al., 2023). In-home sensor monitoring offers a promising approach for this population by enabling the remote collection of functional measures, detection of acute injuries, and tracking of longitudinal health trends (Aramendi et al., 2018; Kim et al., 2017). The increased fidelity of health data supports earlier, targeted interventions for chronic disease management (de Batlle et al., 2021), which could improve the quality of life in remote areas (Janjua et al., 2021).

The impact of remote sensor health monitoring coupled with personalized, self-management intervention (SM) was compared to a standard health education (HE) control cohort in a two-arm randomized clinical trial. As part of the clinical trial, clinicians documented participant encounters using the SOAP framework (Subjective, Objective, Assessment, and Planning) (Weed et al., 1968), which includes narrative and structured content as well as adverse event (AE) occurrences defined by NIA guidelines (of Health and Services, 2024). The notes contain diagnostic and prognostic information about patient conditions, making them ideal features for predicting health events with machine learning. However, since patient encounters occur infrequently, the day-to-day patterns of participant behavior and function are not captured (Filipow et al., 2023). As such, in-home sensor health parameters offer a complementary method for remotely assessing daily physiological functioning with higher frequency measurements.

Classification models for AE and fall injury events were built using BERT-based language model embeddings of semi-structured SOAP notes and in-home sensor measurements, separately and as fused feature sets. BERT language models allow for extracting clinical contexts from semi-structured notes (Arzideh et al., 2025). By integrating the two data streams, we hypothesized that classifier models will be better able to detect signals related to AEs and fall risk. AE predictive models built with integrated clinical and time-series features have the potential to support proactive healthcare by alerting clinicians to emerging problems (Shukla and Marlin, 2021). Such early detection would enable more timely, personalized interventions to prevent hospitalizations or complications, especially in rural communities with limited healthcare access (Esquivel et al., 2023; Ge et al., 2023). This framework for data fusion and predictive analytics holds promise not only for AE detection but also for a wide range of health monitoring applications in aging and chronically ill populations.

## 2. Materials and Methods

### 2.1. Parent Study

A pragmatic two-arm randomized clinical trial was conducted at the University of Missouri Department of Occupational Therapy to compare a self-management intervention to a health education control (Proffitt et al., 2024). All participants had a remote sensor system installed in their home for a one year observation period. Participants randomized to receive the self-management intervention were also encouraged to review their sensor system data and study interveners integrated the data into treatment planning. Participants were recruited from rural areas in central Missouri with a 2020 census block having fewer than 1000 residents per square mile. Study inclusion criteria required participants to be over the age of 65 having difficulty with two or more activities of daily living (ADL)(Katz, 1983), ability to stand upright, internet access, and a trusted support person. 57 participants were included in our analysis across the two study arms as shown in Table **1**. The self-management intervention applied to the intervention cohort utilized the 5A Behavioral Change Model (Ask, Advise, Assess, Assist, and Arrange)(Glasgow et al., 2002) to guide health goals for participants as part of a personalized health plan developed and implemented by the interdisciplinary (registered nurse, occupational therapist, and social worker) team of clinicians. The control cohort were provided with basic health education information and resource aid for treating conditions without guided planning. Patient encounter documentation and instruments were stored in a secure REDCap research data repository (Harris et al., 2009). Clinician encounter notes ensured the integrity of data collected (of Health, 2016), while clinical outcome measures provide the basis for evaluating intervention effectiveness over the course of the clinical trial.

**Table 1:** Participant demographic characteristics at time of enrollment by study arm.

|  | Health Education | Self-Management | Total |
| --- | --- | --- | --- |
| **Sample Size** | 32 | 25 | 57 |
| **Age (years)** |  |  |  |
| Mean (SD) | 75.1 (5.7) | 76.4 (5.8) | 75.7 (5.7) |
| Range | 67–91 | 66–87 | 66–91 |
| **Sex** |  |  |  |
| Male | 13 (40.6%) | 8 (32%) | 21 (46.8%) |
| Female | 19 (59.4%) | 17 (68%) | 36 (63.2%) |
| **Race** |  |  |  |
| White | 32 (100%) | 24 (96%) | 56 (98.2%) |
| Multiple | 0 (0%) | 1 (4%) | 1 (1.8%) |
| **Ethnicity** |  |  |  |
| Hispanic | 0 (0%) | 0 (0%) | 0 (0%) |
| Non-Hispanic | 32 (100%) | 25 (100%) | 57 (100%) |
| **Annual Income** |  |  |  |
| 19–49k | 12 (37.5%) | 14 (56%) | 26 (45.6%) |
| 49–89k | 8 (25%) | 8 (32%) | 16 (28.1%) |
| >90k | 12 (37.5%) | 3 (12%) | 15 (26.3%) |
| **Have Tested Covid-19 Positive** |  |  |  |
| Yes | 22 (68.8%) | 6 (24%) | 28 (49.1%) |
| No | 10 (31.2%) | 19 (76%) | 29 (50.9%) |
| **Healthcare Utilization in Previous 3 Months** |  |  |  |
| Primary | 20 (62.5%) | 22 (88%) | 42 (73.7%) |
| Specialist | 21 (65.6%) | 20 (80%) | 41 (71.9%) |
| ER | 6 (18.8%) | 4 (16%) | 10 (17.5%) |
| Hospital | 2 (6.3%) | 2 (8%) | 4 (7%) |
| Surgery | 5 (15.6%) | 3 (12%) | 8 (14%) |
| Fall | 10 (31.3%) | 8 (32%) | 18 (31.6%) |

### 2.2. Semi-Structured SOAP Notes

Clinician SOAP notes were entered at quarterly follow up sessions and during ad hoc participant calls conducted as a human subjects safety protocol. Attributes for type of AE with categories for death, medical-related, participant complaint, study violation, and other, were applied as binary labels to each SOAP note record. The AE attributes for medical-related and other were further sub-categorized by manual review into fall and non-fall event labels. Fall event labels were applied to records where fall-related injuries were sustained by participants, either as self-reported or detected by a depth sensor. SOAP note contexts were extracted as features and the binary AE markers serve as classification labels for targeting AE occurrence and fall injury events, described in Table **2**. In addition to classification of whether an AE occurred or not (57 samples in controls, 50 intervention samples), a set of models were built for classifying fall injuries (28 samples in controls, 29 intervention samples). There were too few samples across both study cohorts in the remaining AE types for death (1), complaint (2), and violation (1) and medical sub-labels for seizure (3), influenza (4), Covid-19 (7), ER visit (26), inpatient (9), outpatient (27), and nursing home intake (3) to conduct multi-label classification.

**Table 2:**
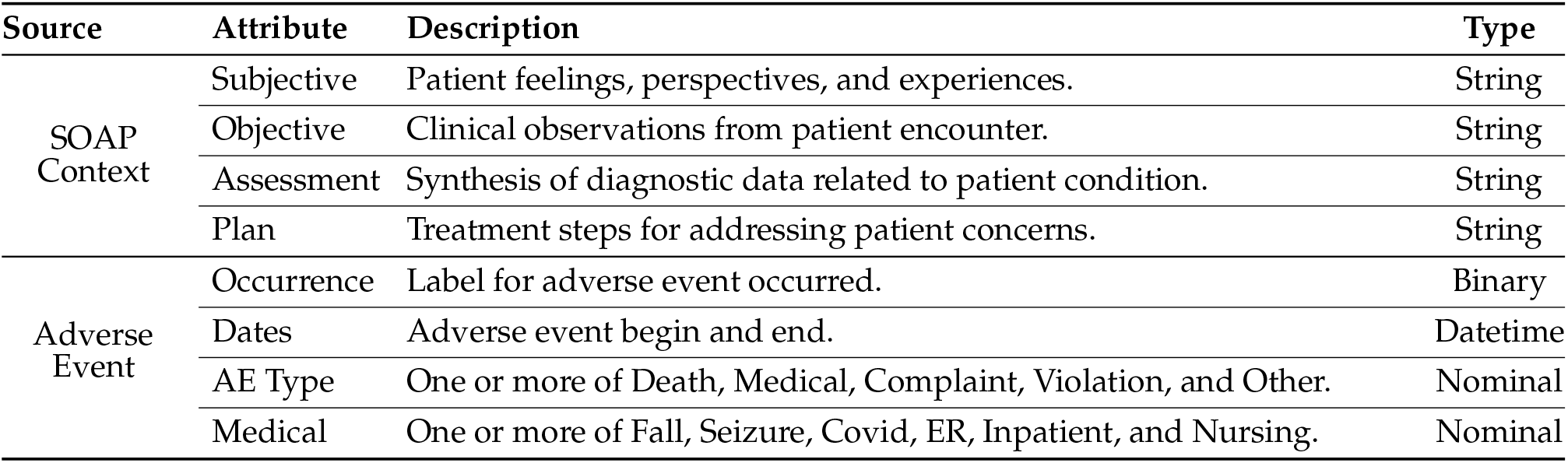
Clinical context and adverse event label types extracted from semi-structured SOAP formatted study documentation notes.

| Source | Attribute | Description | Type |
| --- | --- | --- | --- |
| SOAP Context | Subjective | Patient feelings, perspectives, and experiences. | String |
|  | Objective | Clinical observations from patient encounter. | String |
|  | Assessment | Synthesis of diagnostic data related to patient condition. | String |
|  | Plan | Treatment steps for addressing patient concerns. | String |
| Adverse Event | Occurrence | Label for adverse event occurred. | Binary |
|  | Dates | Adverse event begin and end. | Datetime |
|  | AE Type | One or more of Death, Medical, Complaint, Violation, and Other. | Nominal |
|  | Medical | One or more of Fall, Seizure, Covid, ER, Inpatient, and Nursing. | Nominal |

### 2.3. Sensor-based Health Parameters

Sensor modality-specific health parameter estimates were collected from networked bed and motion systems installed within participant homes, described in Table **4**. The bed mat sensor containing hydraulic pressure transducers beneath the mattress captured ballistocardiogram signals while the participant was in bed, which were deconvolved for pulse and respiration rates and sleep restlessness (Heise et al., 2011). Passive infrared (PIR) motion sensors were installed throughout the home for measuring room level activity aggregated to time interval densities (Wang et al., 2012). Additionally, a depth sensor monitored for fall injuries using an object and floor plane segmentation process and sent logged email notifications when a fall was detected (Stone and Skubic, 2014), which served as a source of labeling for fall injury events. The sensor systems have been deployed and validated at independent living communities and in private residences through research studies funded by the National Institutes of Health and National Library of Medicine (Jain et al., 2020; Rantz et al., 2015).

**Table 3:**
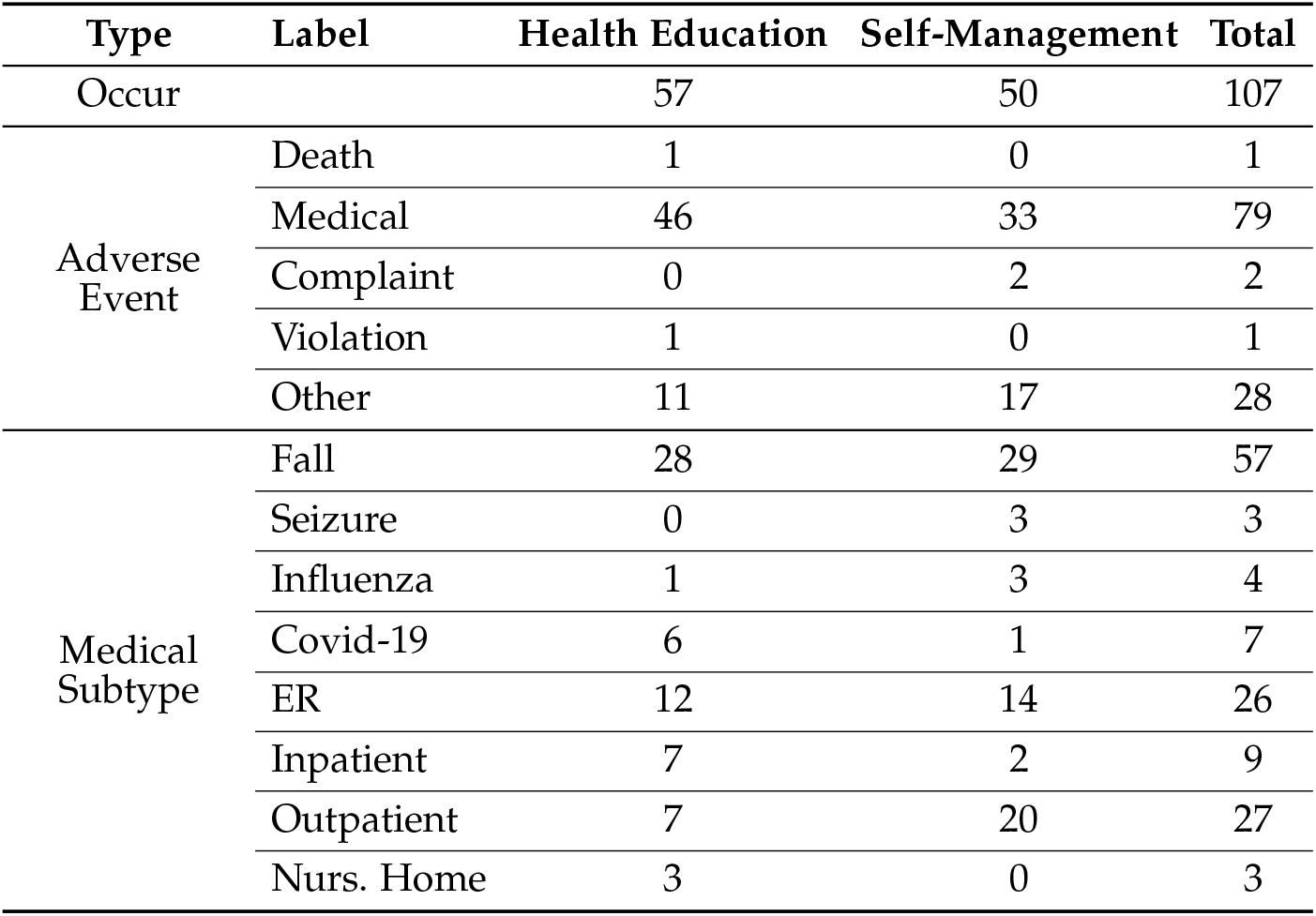
Summary of adverse event occurrence and medical subtype by study arm. Medical event labels are non-exclusive.

**Table 4:**
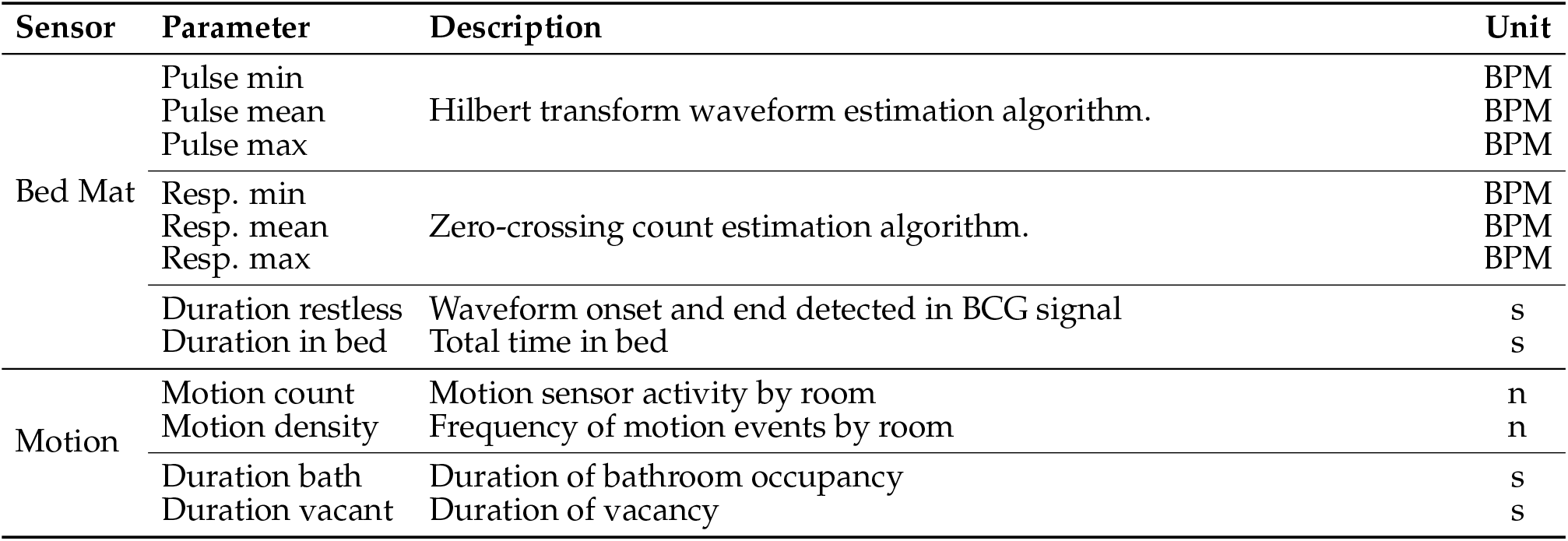
Health parameter estimates and corresponding units as measured by in-home sensor systems. Abbreviations: BCG, ballistocardiography; BPM, beats per minute; s, seconds; in, inches; n, count.

Due to the heterogeneous nature of study participant home environments such as internet and power availability for sensor systems and changes in participant behavior, such as an adjustment in the sleeping area or extended absence from the home, missing values were introduced into sensor data streams. Since data gaps may be representative of behavioral and functional health states, missing values were not removed or imputed prior to model fitting.

### 2.4. Language Model Embeddings

SOAP note component texts were reduced to named-entity recognition (NER) tokens generated by first applying lemmatizer and stemmer functions with the *Natural Language Toolkit* (NLTK) Python library (Bird et al., 2009) prior to fitting them with the ScispaCy *en core sci scibert* biomedical text processing model pipeline (Neumann et al., 2019) and UMLS corpus linker. Named entities were filtered to terms matching UMLS concepts in order to optimize the medical relevancy of the resulting tokens. Language model embeddings were generated using the process outlined in Algorithm 1. Three Bidirectional Encoder Representations from Transformers (BERT) based models were evaluated using the HuggingFace API for BioClinicalBERT (Alsentzer et al., 2019), BioBERT (Lee et al., 2020), and Google’s large BERT-Uncased (Devlin et al., 2019). BioClinicalBERT and BioBERT model embeddings were specifically chosen as they were pretrained on clinical text documents. BioBERT was initialized on the BERT base model and pretrained on PubMed abstracts and the full-text of articles available from PubMedCentral prior to fine-tuning for biomedical tasks. The BioClinicalBERT model had been initialized on the BioBERT language model and pretrained using clinical notes from the MIMIC III notes database (Johnson et al., 2016). The BERT-Uncased model provides a comparative baseline measure against the BioBERT and BioClinicalBERT language model embeddings. Each of the evaluated models mapped the NER tokens matching UMLS terms into a 768-dimensional dense vector space.

#### Algorithm 1: Pseudo code of SOAP note embeddings

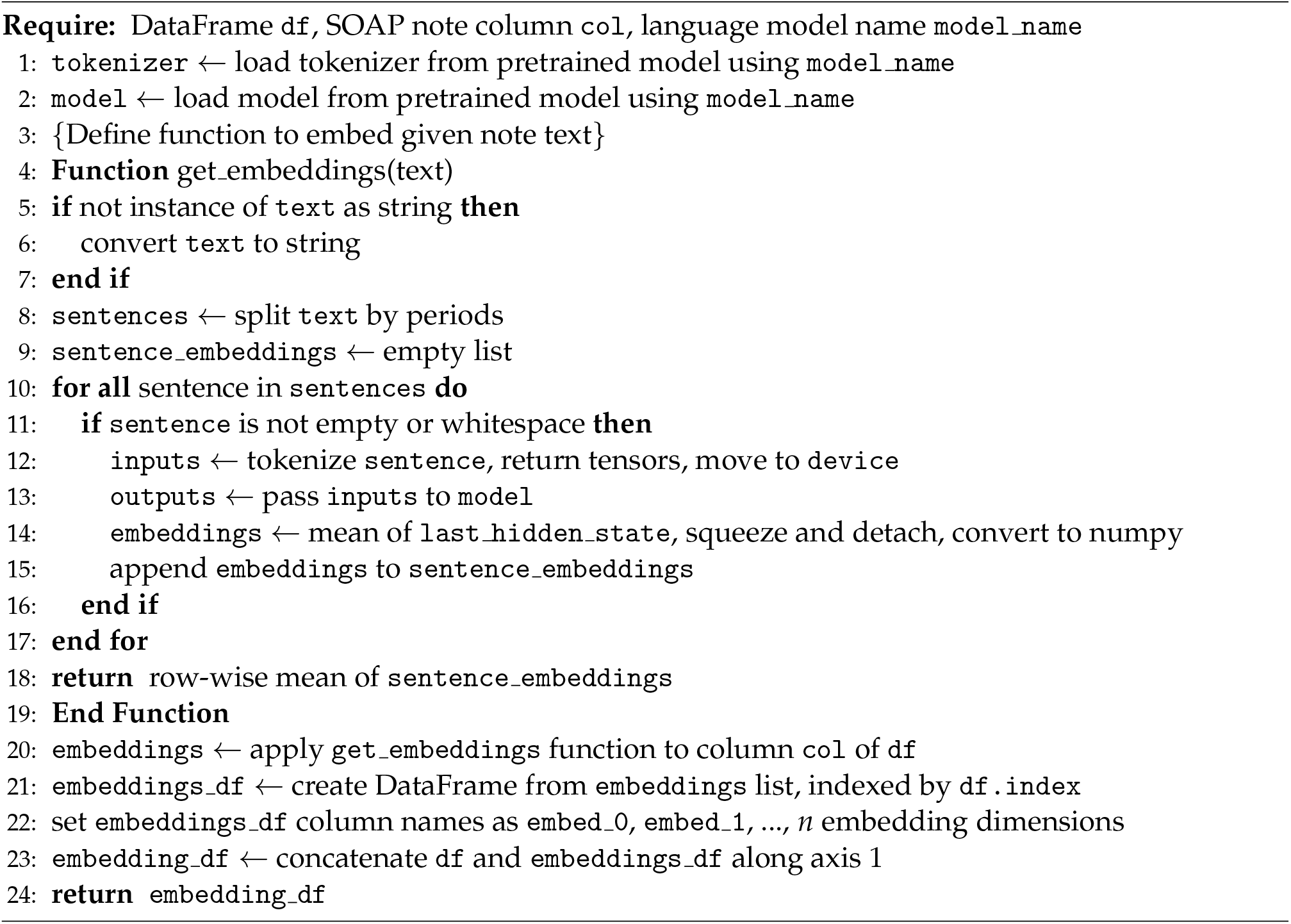

### 2.5. Sensor and SOAP Embeddings Feature Fusion

The sensor-based health parameters were indexed by timestamp and study participant identifiers as continuous time series values during data collection and tabulated, as illustrated in Figure **1**. Summary statistics were then applied for minimum, mean, and maximum of each parameter as daily 24-hour aggregations. The daily aggregations were then passed through Min-Max normalization and segmented to the 30 days preceding the date of each clinical note entered for each participant identifier, shown in Figure **2**. The 30-day segmented sensor health features leading to an AE were merged with clinical text language model embeddings using backfill interpolation on the low frequency embeddings by date. The fused feature sets were then labeled row-wise with binary event markers for AE occurrence and fall injury events identified from the original SOAP notes.

**Fig. 1:**
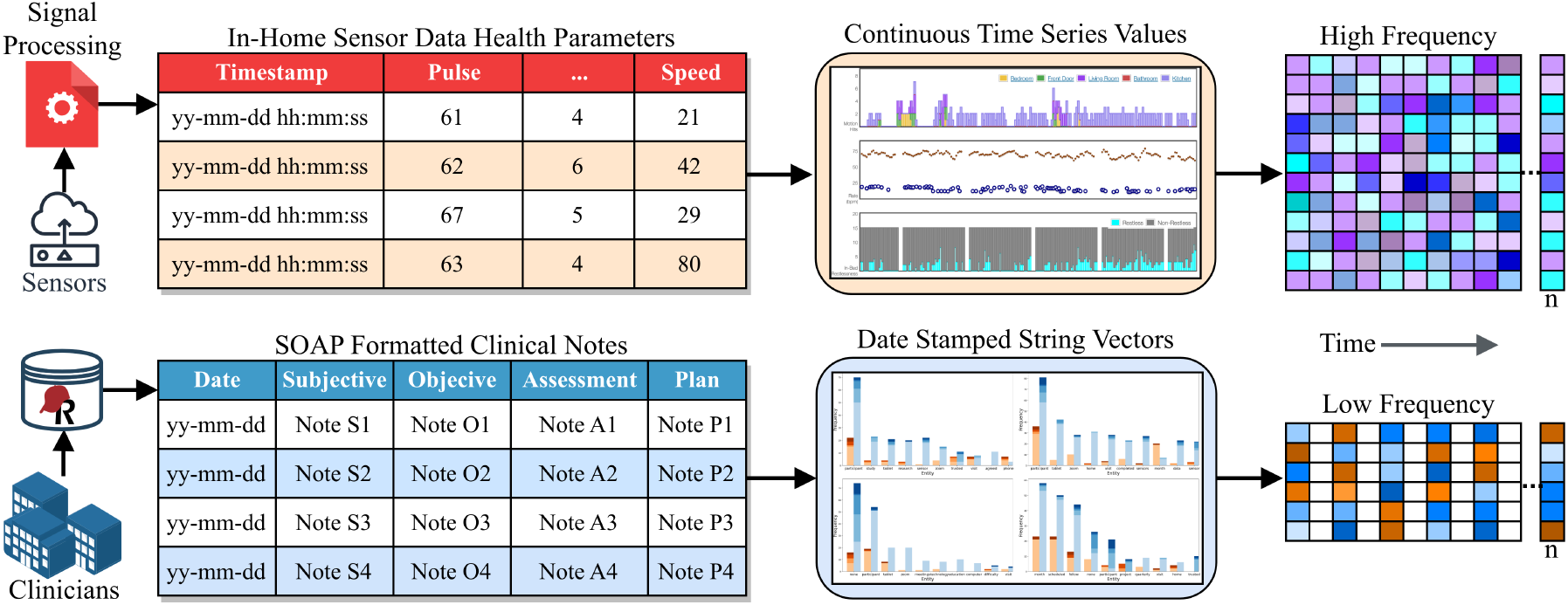
Data collection and aggregation of time-series sensor data and dated SOAP clinical notes.

**Fig. 2:**
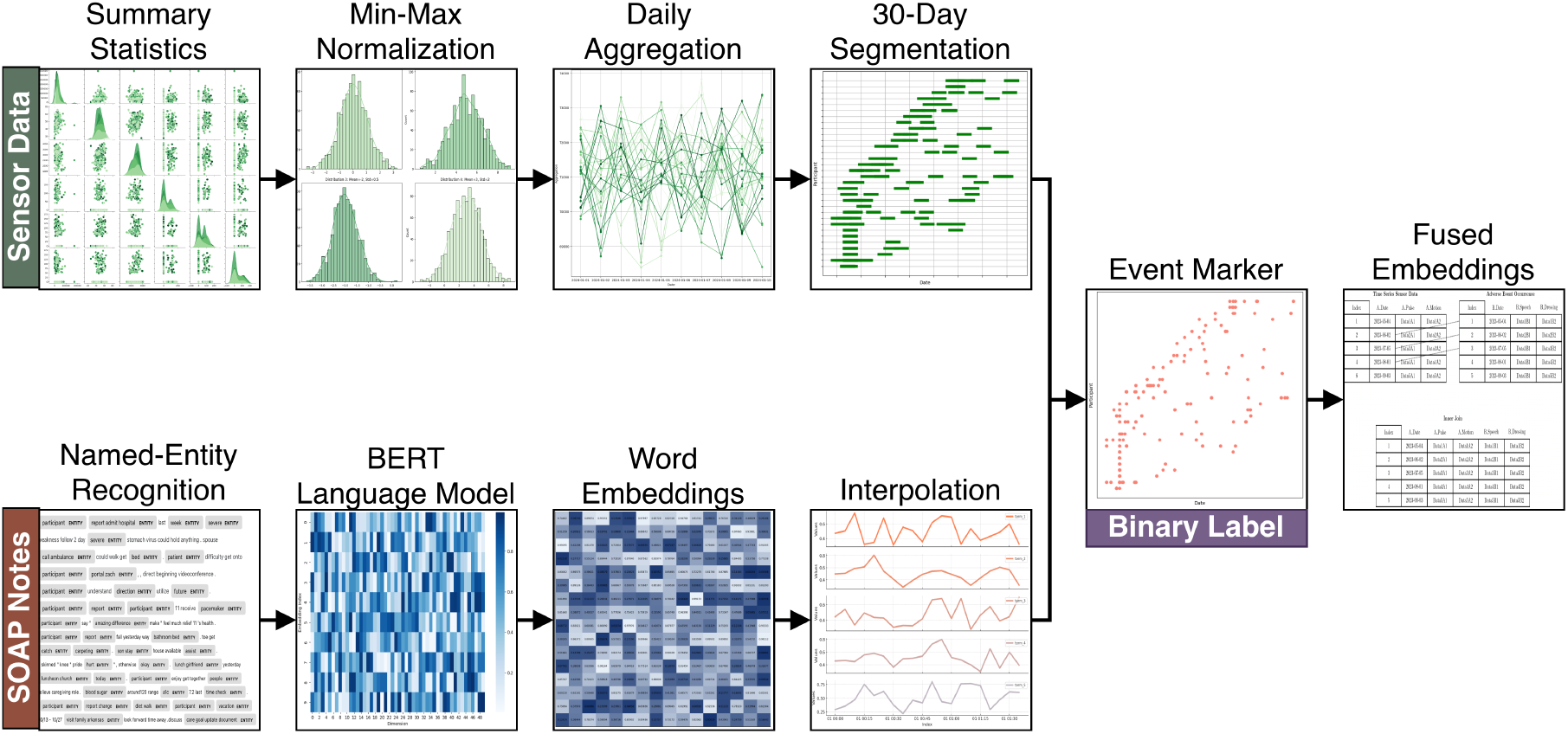
Preprocessing of sensor-based health parameters and SOAP note embeddings.

### 2.6. Classification Models with Iterative Feature Selection

Models targeting AE occurrence and fall injury events were fit on clinical text embeddings, and sensor health features, and on fused predictor sets of combined sensor features with text embeddings. The embedding language model features for individual SOAP note components were evaluated independent of one another to determine how clinical note perspectives associate with AEs.

All classifiers were built using XGBoost Python library’s *XGBClassifier* (Chen and Guestrin, 2016) with hyperparameters tuned with *GridSearchCV* model selection search routine in the scikit-learn Python library (Pedregosa et al., 2011), as illustrated in Figure **3**. XGboost was chosen through preliminary testing in computation times and accuracy metrics over other gradient boosting algorithms and for its robust handling of null values in sensor feature streams.

**Fig. 3:**
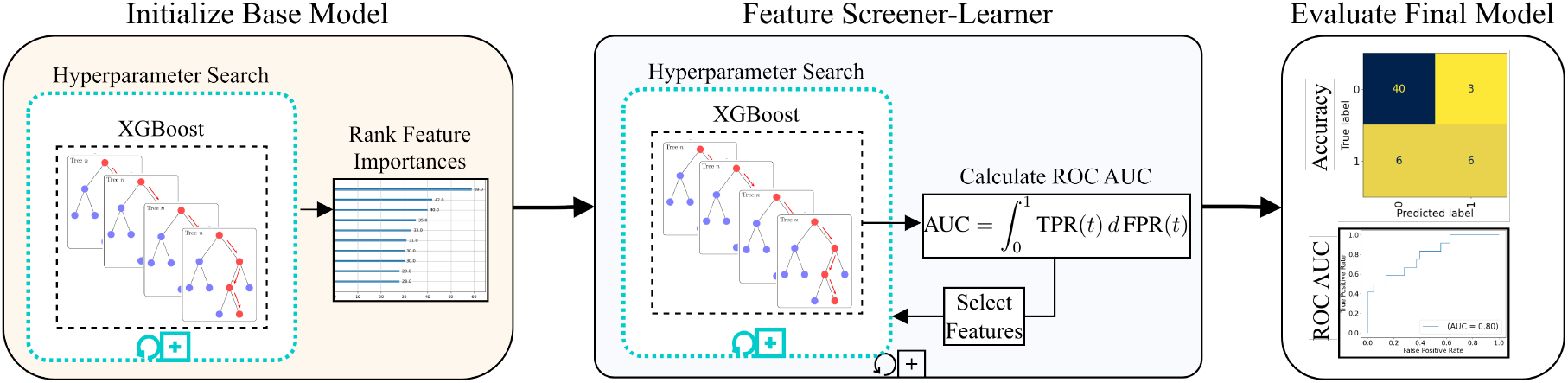
XGBoost classification model pipeline using iterative feature screener-learner.

### 2.7. Model Performance Evaluation Metrics

We evaluated model performance using the metrics defined in Equations 1-6. In our classification tasks, accuracy (Eq:1) serves as a measure of overall correctness by taking the ratio between the true positive and true negative observations to the total observations. Recall (Eq:2) is the ratio of true positives to the total number of actual positive observations as a measure of the model’s ability to correctly identify positive instances. Precision (Eq:3) is the proportion of correct classified positives to the total number of classified as positive. AUROC (Area Under the Receiver Operating Characteristic Curve) (Eq:5) evaluates the ability of classifier (*c*) to differentiate between classes, with a value closer to 1 indicating better performance. The AUROC Confidence Interval (Eq:6) indicates the range of values within which the true AUROC likely falls as an estimate of the variability in the AUROC value.

Performance metrics were calculated using a training-test hold out of 80% and 20%, respectively, from the observations sets. The area under the ROC curve (AUROC) serves as our primary metric followed by classification accuracy due to the detrimental effect that misclassified AEs have on the health of participants. The scikit-learn Python library was used for metric calculations on the test predictions from the tuned model returned by *GridSearchCV* with *sklearn*.*metrics* score functions for accuracy, precision, recall, F1, and AUROC using 5-fold cross validation and predicted class probabilities.

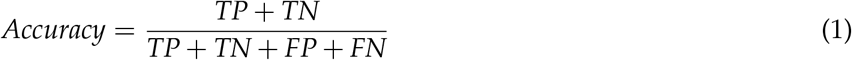

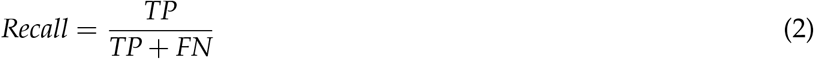

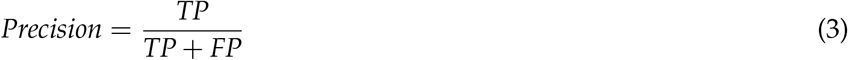

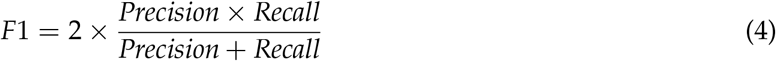

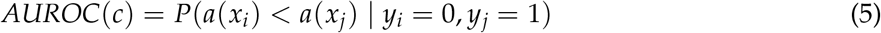

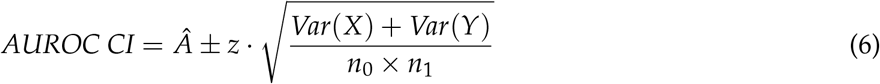

## 3. Results

### 3.1. Comparison of Structured Clinical Content by Event Type

SOAP notes extracted as adverse event (AE) related and non-adverse event (non-AE) related were compared across HE controls and SM intervention cohorts to determine if there are differences in the number of notes (observations) and the number of NER tokens (words) between them after filtering entities against the UMLS corpus. In the HE cohort, plotted in Figure **4**, there were fewer AE notes than non-AE notes across all SOAP sections. For AEs, the number of observations ranged from 37 (Assessment) to 61 (Objective), while non-AEs ranged from 54 (Assessment) to 131 (Objective). Token counts followed a similar pattern, with AE notes having fewer tokens per section (Subjective: 167, Objective: 287, Assessment: 85, Plan: 101) compared to non-AE notes (Subjective: 305, Objective: 584, Assessment: 160, Plan: 291). The number of unique tokens per section was also lower in AE notes. The SM intervention cohort, Figure **5**, had a higher total number of notes. For AE notes, observations per section ranged from 84 (Assessment) to 95 (Subjective), and for non-AE notes from 312 (Assessment) to 340 (Objective). The AE notes contained fewer tokens in the Objective, Assessment, and Plan sections compared to non-AEs, but unlike the HE controls, there were more tokens in the Subjective section (AE: 1003; non-AE: 2920). Unique token counts were higher in non-AE notes across all sections. Additionally, AE notes were consistently shorter having fewer observations at a lower count of tokens compared to non-AE note across SOAP note sections, with the largest differences between the Objective and Subjective sections. The SM cohort demonstrated higher documentation rates but demonstrated the same trend of shorter AE notes.

**Fig. 4:**
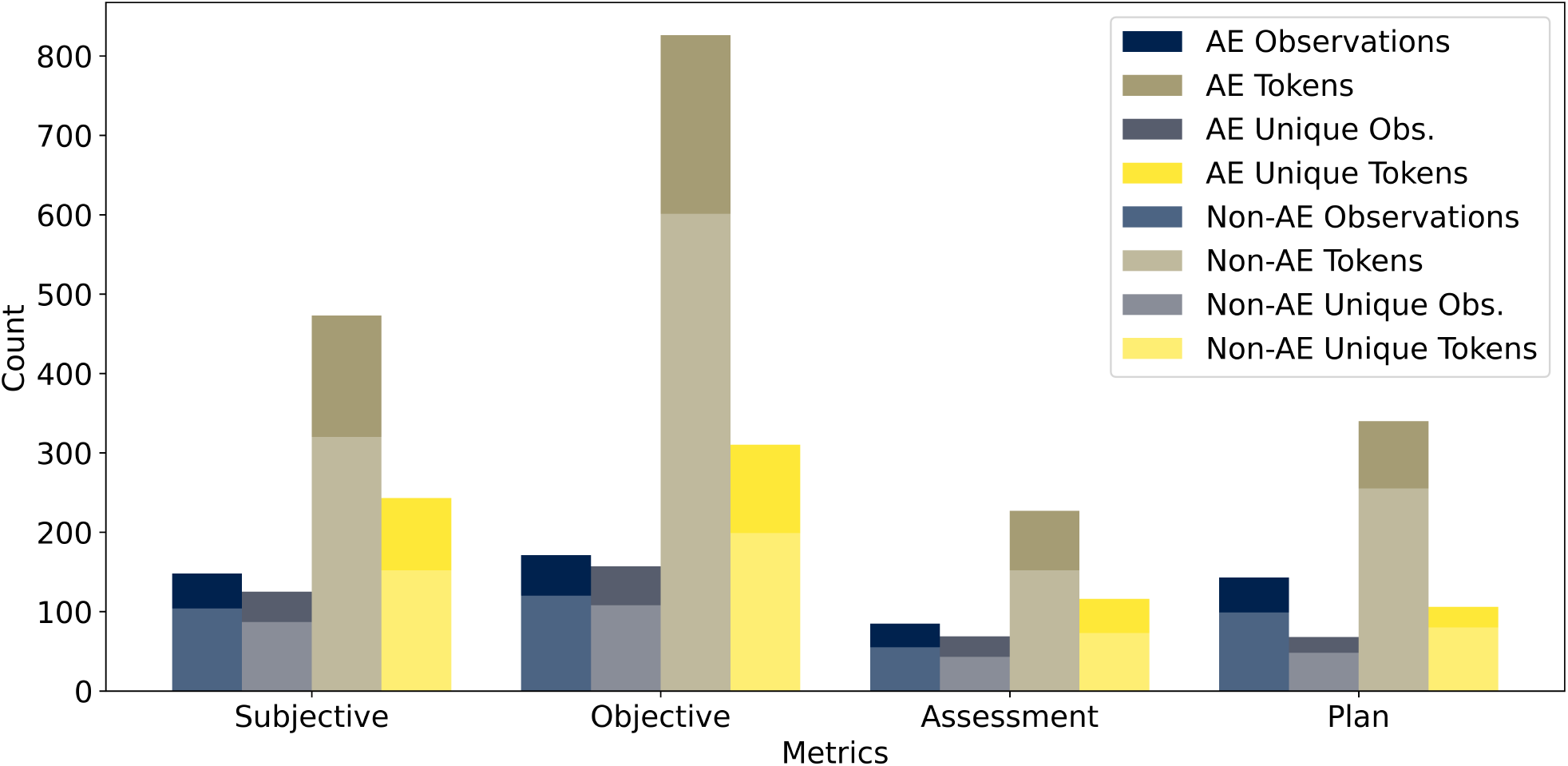
Named-entity tokens across HE SOAP Notes by adverse event occurrence. The increased quantity of objective notes for the control arm is consistent with study protocol in that participants are not receiving planning intervention and less queried regarding changes in health.

**Fig. 5:**
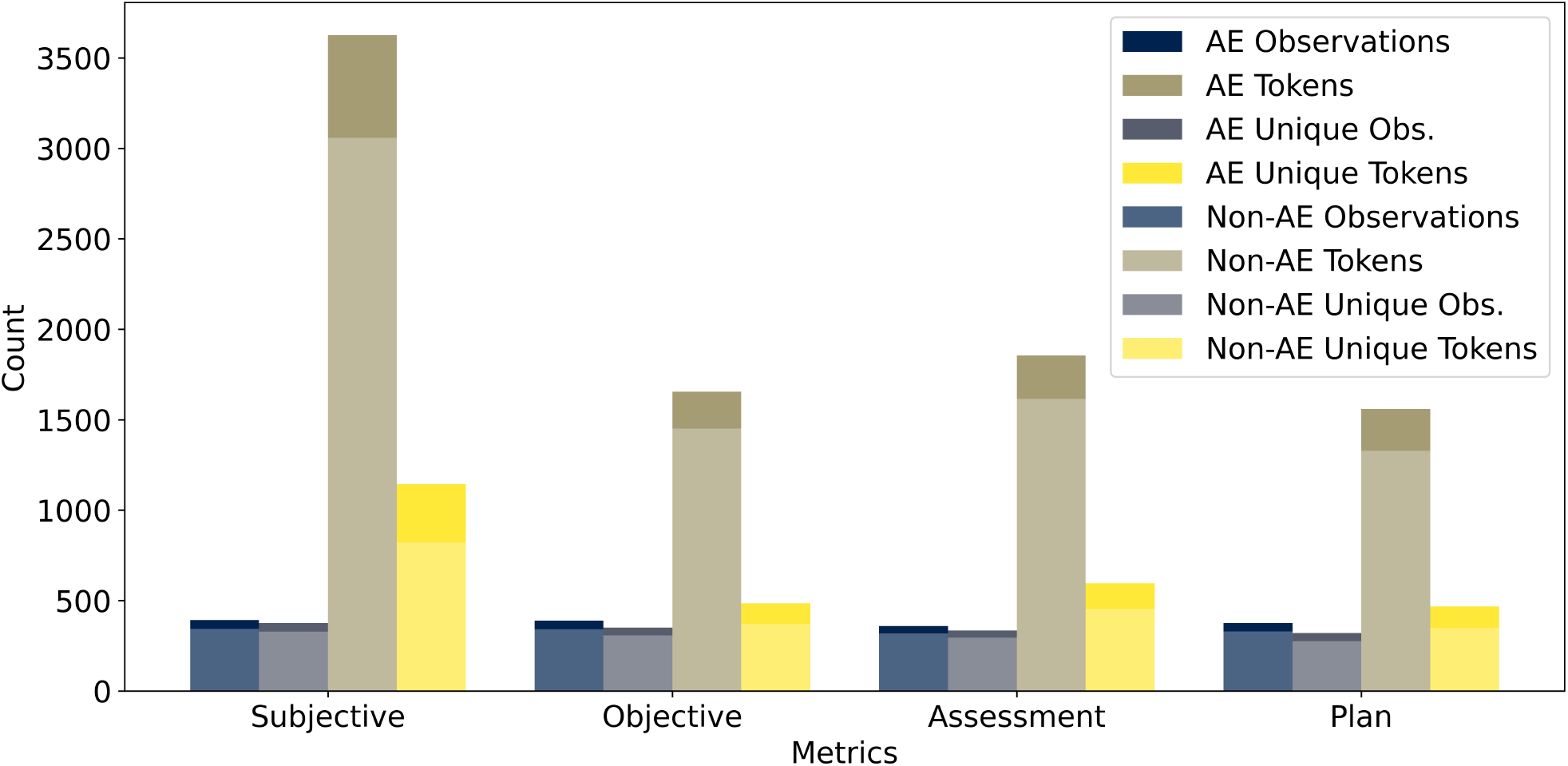
Named-entity tokens across SM SOAP Notes by adverse event occurrence. The intervention group contains higher number of subjective notes and note tokens, indicating that clinicians have more engaged with capturing participant contexts in the documentation.

### 3.2. Adverse Event Occurrence Classifiers

Model performance metrics were compared across control (HE) and intervention (SM) study cohorts for AE and fall injury event classification. Metrics are reported for models fit on individual language model embeddings and sensor features, in Table **6**, and sensor features fused with language model embeddings, in Table **7**. These comparisons evaluate whether the differing clinical context and perspectives in SOAP note components affect model performance by study arm. Additionally, we evaluate whether fusing data from multiple modalities enhances the predictive accuracy of classification models. Classifier model performance metrics for predicting the occurrence of AEs are shown beneath the *AE Occurrence* section of Table **6** and Table **7**. Performances across embedding models and sensor features are summarized below, with a focus on AUROC and recall. The 95% CI for AUROC was used to assess significance between models and feature sets.

**Table 5:**
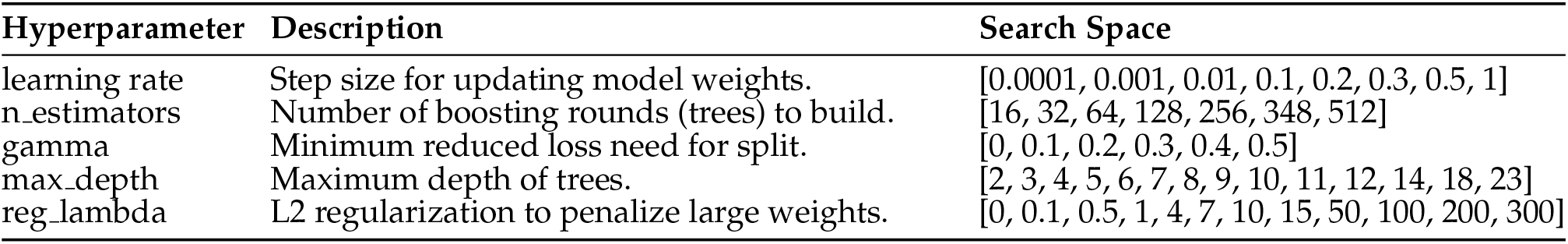
XGBoost algorithm hyperparameters searched during learner tuning.

**Table 6:**
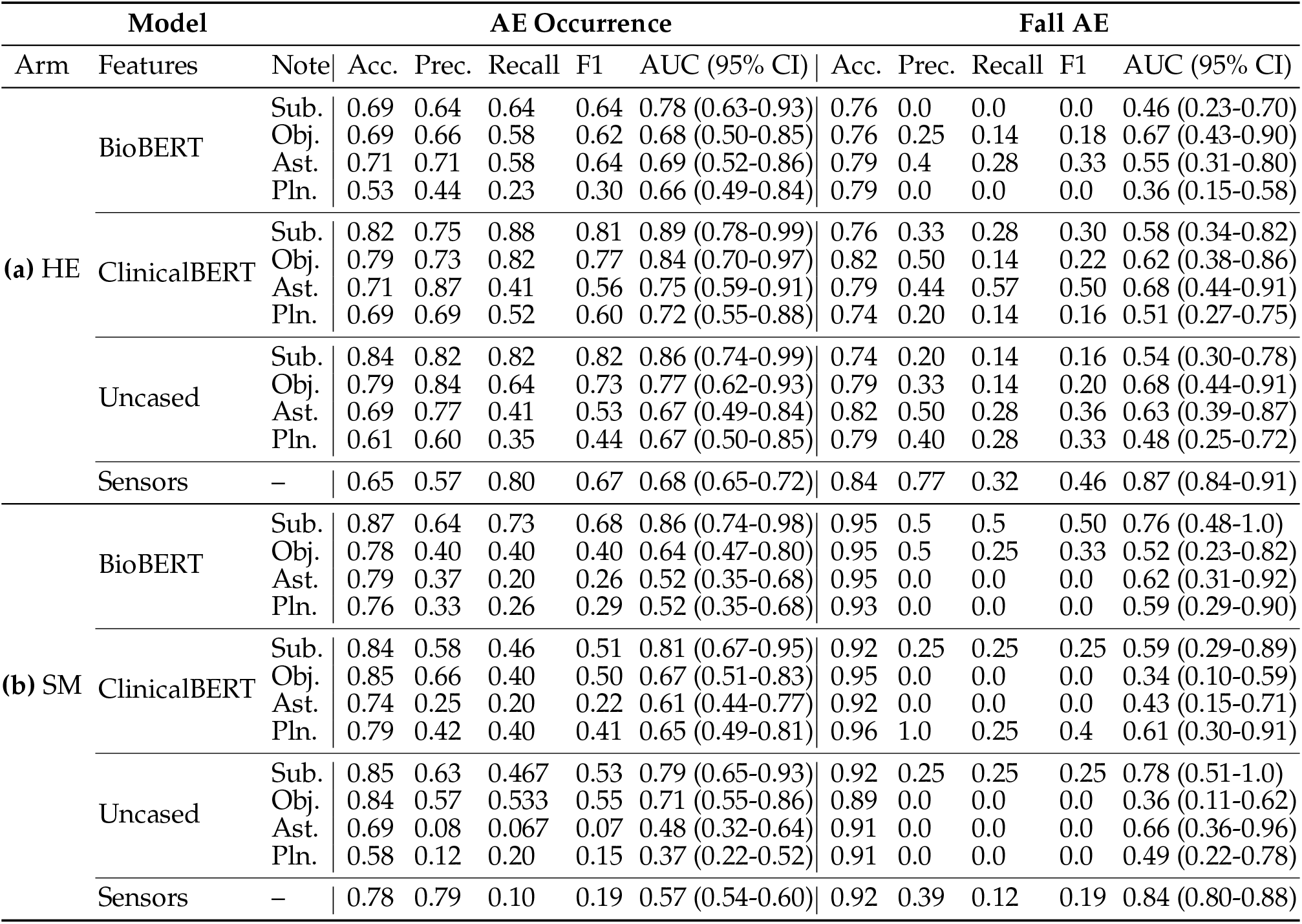
Embedding model performance metrics for AE occurrence by **(a)** HE (control) and **(b)** SM (intervention) study arms.

**Table 7:** Embedding model performance metrics for AE occurrence by **(a)** HE (control) and **(b)** SM (intervention) study arms.

| Model |  |  | AE Occurrence |  |  |  |  | Fall AE |  |  |  |  |
| --- | --- | --- | --- | --- | --- | --- | --- | --- | --- | --- | --- | --- |
| Arm | Features | Note | Acc. | Prec. | Recall | F1 | AUC (95% CI) | Acc. | Prec. | Recall | F1 | AUC (95% CI) |
| (a) HE | BioBERT & Sensors | Sub. | 1.0 | 1.0 | 1.0 | 1.0 | 1.0 (1.0) | 1.0 | 1.0 | 1.0 | 1.0 | 1.0 (1.0) |
|  |  | Obj. | 0.99 | 0.99 | 1.0 | 0.99 | 1.0 (0.99–1.0) | 1.0 | 1.0 | 1.0 | 1.0 | 1.0 (1.0) |
|  |  | Ast. | 0.83 | 1.0 | 0.63 | 0.77 | 0.89 (0.86–0.91) | 0.94 | 0.99 | 0.74 | 0.85 | 0.92 (0.9–0.95) |
|  |  | Pln. | 0.94 | 0.93 | 0.93 | 0.93 | 0.98 (0.98–0.99) | 0.95 | 0.90 | 0.85 | 0.87 | 0.98 (0.97–0.99) |
|  | ClinicalBERT & Sensors | Sub. | 0.99 | 0.99 | 1.0 | 0.99 | 1.0 (0.99–1.0) | 1.0 | 1.0 | 1.0 | 1.0 | 1.0 (1.0) |
|  |  | Obj. | 1.0 | 1.0 | 1.0 | 1.0 | 1.0 (1.0) | 1.0 | 1.0 | 1.0 | 1.0 | 1.0 (1.0) |
|  |  | Ast. | 0.83 | 0.99 | 0.63 | 0.77 | 0.89 (0.86–0.91) | 0.94 | 1.0 | 0.74 | 0.85 | 0.92 (0.90–0.95) |
|  |  | Pln. | 0.93 | 0.92 | 0.93 | 0.93 | 0.98 (0.98–0.99) | 0.95 | 0.4 | 0.85 | 0.87 | 0.97 (0.96–0.99) |
|  | Uncased & Sensors | Sub. | 1.0 | 1.0 | 1.0 | 1.0 | 1.0 (1.0) | 1.0 | 1.0 | 1.0 | 1.0 | 1.0 (1.0) |
|  |  | Obj. | 1.0 | 1.0 | 1.0 | 1.0 | 1.0 (1.0) | 1.0 | 1.0 | 1.0 | 1.0 | 1.0 (1.0) |
|  |  | Ast. | 0.83 | 0.99 | 0.63 | 0.77 | 0.89 (0.86–0.91) | 0.94 | 1.0 | 0.74 | 0.85 | 0.93 (0.90–0.95) |
|  |  | Pln. | 0.93 | 0.92 | 0.93 | 0.93 | 0.98 (0.98–0.99) | 0.90 | 1.0 | 0.55 | 0.71 | 0.96 (0.93–0.98) |
| (b) SM | BioBERT & Sensors | Sub. | 1.0 | 1.0 | 1.0 | 1.0 | 1.0 (1.0) | 1.0 | 1.0 | 1.0 | 1.0 | 1.0 (1.0) |
|  |  | Obj. | 0.99 | 1.0 | 0.99 | 0.99 | 1.0 (0.99–1.0) | 1.0 | 1.0 | 1.0 | 1.0 | 1.0 (1.0) |
|  |  | Ast. | 0.98 | 1.0 | 0.93 | 0.96 | 0.99 (0.99–1.0) | 0.99 | 1.0 | 0.87 | 0.93 | 0.99 (0.99–1.0) |
|  |  | Pln. | 0.98 | 0.99 | 0.95 | 0.97 | 1.0 (0.99–1.0) | 1.0 | 1.0 | 1.0 | 1.0 | 1.0 (1.0) |
|  | ClinicalBERT & Sensors | Sub. | 1.0 | 1.0 | 1.0 | 1.0 | 1.0 (1.0) | 1.0 | 1.0 | 1.0 | 1.0 | 1.0 (1.0) |
|  |  | Obj. | 0.99 | 1.0 | 0.99 | 0.99 | 1.0 (0.99–1.0) | 1.0 | 1.0 | 1.0 | 1.0 | 1.0 (1.0) |
|  |  | Ast. | 0.98 | 1.0 | 0.93 | 0.96 | 0.99 (0.99–1.0) | 0.99 | 1.0 | 0.87 | 0.93 | 0.99 (0.99–1.0) |
|  |  | Pln. | 0.99 | 1.0 | 0.96 | 0.98 | 1.0 (0.99–1.0) | 0.91 | 0.0 | 0.0 | 0.0 | 0.52 (0.47–0.57) |
|  | Uncased & Sensors | Sub. | 1.0 | 1.0 | 1.0 | 1.0 | 1.0 (1.0) | 1.0 | 1.0 | 1.0 | 1.0 | 1.0 (1.0) |
|  |  | Obj. | 0.99 | 1.0 | 0.99 | 0.99 | 1.0 (0.99–1.0) | 1.0 | 1.0 | 1.0 | 1.0 | 1.0 (1.0) |
|  |  | Ast. | 0.98 | 1.0 | 0.93 | 0.96 | 0.99 (0.99–1.0) | 0.99 | 1.0 | 0.87 | 0.93 | 0.99 (0.99–1.0) |
|  |  | Pln. | 0.99 | 1.0 | 0.96 | 0.98 | 1.0 (0.99–1.0) | 1.0 | 1.0 | 1.0 | 1.0 | 1.0 (1.0) |

#### 3.2.1. Non-Fused Sensor and SOAP Embeddings Features

For the HE control arm, reported in the *AE Occurrence* column group in Table **6(a)**, BioBERT had the highest AUROC and moderate recall when fit on Subjective notes (AUROC=0.78(0.63–0.93), Recall=0.64), followed by Assessment (AUROC=0.69(0.52–0.86), Recall=0.58), Objective (AUROC=0.68(0.50–0.85), Recall=0.58), and Planning (AUROC=0.66(0.49–0.84), Recall=0.23). The confidence intervals for AUROC overlapped across all SOAP sections, indicating no significant differences among BioBERT models. ClinicalBERT performed best on Subjective notes (AUROC=0.89(0.78–1.0), Recall=0.88), followed by Objective (AUROC=0.84(0.70–0.97), Recall=0.82), Assessment (AUROC=0.75(0.59–0.91), Recall=0.41), and Planning (AUROC=0.72(0.55–0.88), Recall=0.52). While the Subjective notes had the highest AUROC, the confidence intervals for BioClinicalBERT models overlapped, so differences were not statistically significant. For the BERT-Uncased model, the highest AUROC was also seen with Subjective notes (AUROC=0.86(0.74–0.99), Recall=0.82), with Objective (AUROC=0.77(0.62–0.93), Recall=0.64), Assessment (AUROC=0.67(0.49–0.84), Recall=0.41), and Planning (AUROC=0.67(0.50–0.85), Recall=0.35) showing lower values. As with the other embeddings, AUROC confidence intervals for BERT-Uncased models didn’t indicate significant differences among SOAP notes. Sensor features alone had an AUROC of 0.68(0.65–0.72), Recall=0.80. The narrow confidence interval for AUROC indicates a stable estimate, but the AUROC was lower than the best-performing embedding models. When comparing across feature sets, both BioClinicalBERT and BERT-Uncased Subjective notes models significantly outperformed sensor features in terms of AUROC, with non-overlapping upper confidence interval bounds. Sensor AUROC overlapped with all BioBERT sections and the lower-performing BioClinicalBERT and BERT-Uncased SOAP sections without significant improvement between them. Sensor features provided higher recall than most embedding models, but at lower AUROC. No statistically significant differences were found among the best SOAP sections within each embedding model.

In the SM intervention study arm, reported in the *AE Occurrence* column group in Table **6(b)**, as in HE controls BioBERT had the highest AUROC when fit on Subjective notes (AUROC=0.86(0.74–0.98), Recall=0.73), with Objective (AUROC=0.64(0.47–0.80), Recall=0.40), Assessment (AUROC=0.52(0.35–0.68), Recall=0.20), and Planning (AUROC=0.52(0.35–0.68), Recall=0.26) sections showing lower AUROCs and recall. The confidence intervals for BioBERT Subjective did not overlap with those for Assessment and Planning, indicating significantly better performance on Subjective notes, though there was some overlap with Objective. ClinicalBERT also performed best on Subjective notes (AUROC=0.81(0.67–0.95), Recall=0.46), followed by Objective (AUROC=0.67(0.51–0.83), Recall=0.40), Planning (AUROC=0.65(0.49–0.81), Recall=0.40), and Assessment (AUROC=0.61(0.44–0.77), Recall=0.20). The AUROC confidence intervals for all BioClinicalBERT SOAP sections overlapped without statistical significance. For the BERT-Uncased model, Subjective notes resulted in the highest AUROC (AUROC=0.79(0.65–0.93), Recall=0.47), followed by Objective (AUROC=0.71(0.55–0.86), Recall=0.53), Assessment (AUROC=0.48(0.32–0.64), Recall=0.07), and Planning (AUROC=0.37(0.22–0.52), Recall=0.20). Subjective and Objective were significantly better compared to Planning, and Subjective was better compared to Assessment notes, as well. Sensor features had an AUROC of 0.57(0.54–0.60) with much lower recall (Recall=0.10) compared to the text embedding models, but had a more stable, narrower confidence interval. When comparing across feature sets, BioBERT and BioClinicalBERT Subjective notes both reached high AUROCs (*>*0.8), with BioBERT having higher recall, which were found to be significantly better compared to BERT-Uncased Assessment and Planning sections and Sensor features.

#### 3.2.2. Fused Sensor and SOAP Embeddings Features

Integrating the sensor data with the language model embeddings resulted in improved model performance for both study arms across the four SOAP clinical note contexts. In the HE control arm, shown in the *AE Occurrence* column group in Table **7a**, fused BioBERT features had perfect AUROC when fit on Subjective notes for AE Occurrence (AUROC=1.0(1.0), Recall=1.0) and Objective notes (AUROC=1.0(0.99–1.0), Recall=1.0). Assessment (AUROC=0.89(0.86–0.91), Recall=0.63) and Planning (AUROC=0.98(0.98–0.99), Recall=0.93) had lower performance, with the confidence interval for Assessment not overlapping with the perfect AUROC of Subjective and Objective sections, indicating significantly worse discrimination for Assessment. The Planning section also performed worse than Subjective and Objective, but with high AUROC and minimal CI overlap at the lower bound. ClinicalBERT features fused with sensors showed very similar patterns, with perfect or near-perfect AUROC for Subjective (AUROC=1.0(0.99–1.0), Recall=1.0) and Objective (AUROC=1.0(1.0), Recall=1.0) sections. Assessment (AUROC=0.89(0.86–0.91), Recall=0.63) was again significantly lower due to non-overlapping CIs, and Planning (AUROC=0.98(0.98–0.99), Recall=0.93) performed slightly worse than Subjective and Objective. BERT-Uncased fused models resulted in perfect AUROC and recall for both Subjective (AUROC=1.0(1.0), Recall=1.0) and Objective (AUROC=1.0(1.0), Recall=1.0) sections. Assessment (AUROC=0.89(0.86–0.91), Recall=0.63) had lower performance, while Planning (AUROC=0.98(0.98–0.99), Recall=0.93) showed near perfect AUROC. For the BERT-Uncased fused models, Assessment and Planning performing significantly worse than the Subjective and Objective sections. Across all feature sets, models fit on Subjective and Objective notes consistently had perfect or near-perfect AUROC, with confidence intervals that do not overlap the lower AUROC values seen in Assessment and (to a lesser degree) Plan. This demonstrates that, for AE Occurrence models in the HE arm, Subjective and Objective sections provided significantly better discrimination than Assessment and Planning when using any embedding model combined with sensor features.

For the SM arm, reported in the *AE Occurrence* column group in Table **7b**, fused BioBERT resulted in perfect AUROC for AE Occurrence when fit on Subjective notes (AUROC=1.0(1.0), Recall=1.0) and Objective notes (AUROC=1.0(0.99–1.0), Recall=0.99). Assessment (AUROC=0.99(0.99–1.0), Recall=0.93) and Plan (AUROC=1.0(0.99–1.0), Recall=0.95) also demonstrated high performance. The confidence intervals for all note sections overlapped, indicating no significant differences among these sections. ClinicalBERT fused feature had the same performances, with Subjective (AUROC=1.0(1.0), Recall=1.0), Objective (AUROC=1.0(0.99–1.0), Recall=0.99), Assessment (AUROC=0.99(0.99–1.0), Recall=0.93), and Plan (AUROC=1.0(0.99–1.0), Recall=0.96) having perfect or near-perfect AUROCs. The confidence intervals again were overlapping, so no significant differences were observed among SOAP sections. Fused BERT-Uncased models also had perfect AUROC and recall for Subjective notes (AUROC=1.0(1.0), Recall=1.0) and Objective notes (AUROC=1.0(0.99–1.0), Recall=0.99), with Assessment (AUROC=0.99(0.99–1.0), Recall=0.93) and Plan (AUROC=1.0(0.99–1.0), Recall=0.96), with overlapping confidence intervals. All fused feature set models for AE Occurrence in the SM intervention arm (BioBERT, BioClinicalBERT, BERT-Uncased) performed at or near perfect across all SOAP sections, and there were no statistically significant differences among them.

### 3.3. Fall Injury Event Classifiers

Models for classifying AEs specific to fall injury were evaluated, displayed in the rightmost Fall AE section of Table **6** and Table **7**. As a secondary label for AE occurrences, it was expected that fall injuries will be more difficult to classify due to having lower number of examples to train models from.

#### 3.3.1. Non-Fused Sensor and SOAP Embeddings Features

In the HE controls, outlined in the *Fall AE* column group in Table **6(a)**, BioBERT had the highest AUROC for Fall AE when fit on Objective notes (AUROC=0.67(0.43–0.90), Recall=0.14), followed by Assessment (AUROC=0.55(0.31–0.80), Recall=0.28), Subjective (AUROC=0.46(0.23–0.70), Recall=0.0), and Planning (AUROC=0.36(0.15–0.58), Recall=0.0). Confidence intervals for AUROC overlapped across all BioBERT SOAP sections, so differences were not statistically significant. ClinicalBERT performed best on Assessment notes for Fall AE (AUROC=0.68(0.44–0.91), Recall=0.57), followed by Objective (AUROC=0.62(0.38–0.86), Recall=0.14), Subjective (AUROC=0.58(0.34–0.82), Recall=0.28), and Planning (AUROC=0.51(0.27–0.75), Recall=0.14). The AUROC confidence intervals across SOAP notes overlapped, indicating no significant differences. The BERT-Uncased model had the highest AUROC for Fall AE with Objective notes (AUROC=0.68(0.44–0.91), Recall=0.14), followed by Assessment (AUROC=0.63(0.39–0.87), Recall=0.28), Subjective (AUROC=0.54(0.30–0.78), Recall=0.14), and Planning (AUROC=0.48(0.25–0.72), Recall=0.28). Again, confidence intervals indicated no significant improvements between SOAP notes. Sensor features alone produced the highest overall AUROC for Fall AE (AUROC=0.87(0.84–0.91), Recall=0.32), with a narrow confidence interval. The AUROC for sensors was notably higher than all text-based models, and the upper bound of the embedding model CIs did not overlap with the lower bound of the sensor features, indicating that sensor features performed significantly better than any embedding-based SOAP section for Fall AE detection in the HE arm. There were no significant differences among SOAP sections across embedding models.

For the SM intervention arm, shown in the *Fall AE* column group in Table **6(b)**, BioBERT had the highest AUROC for Fall AE in the SM arm when fit on Subjective notes (AUROC=0.76(0.48–1.0), Recall=0.5), followed by Objective (AUROC=0.52(0.23–0.82), Recall=0.25), Assessment (AUROC=0.62(0.31–0.92), Recall=0.0), and Planning (AUROC=0.59(0.29–0.90), Recall=0.0). The AUROC confidence intervals for all BioBERT SOAP sections overlapped, indicating no significant differences among them. ClinicalBERT performed best for Fall AE on Planning notes (AUROC=0.61(0.30–0.91), Recall=0.25), followed by Subjective (AUROC=0.59(0.29–0.89), Recall=0.25), Assessment (AUROC=0.43(0.15–0.71), Recall=0.0), and Objective (AUROC=0.34(0.10–0.59), Recall=0.0). All confidence intervals overlapped, so no significant differences were observed among BioClinical-BERT SOAP sections. The BERT-Uncased model had the highest AUROC for Fall AE with Subjective notes (AUROC=0.78(0.51–1.0), Recall=0.25), followed by Assessment (AUROC=0.66(0.36–0.96), Recall=0.0), Planning (AUROC=0.49(0.22–0.78), Recall=0.0), and Objective (AUROC=0.36(0.11–0.62), Recall=0.0). Overlapping AUROC confidence intervals among BERT-Uncased SOAP sections indicate no significant differences. Sensor features had the highest overall AUROC for Fall AE (AUROC=0.84(0.80–0.88), Recall=0.12), with a narrow confidence interval. Sensor features performed significantly better than the Objective notes for every embedding model, as well as the BioClinicalBERT Assessment and BERT-Uncased Planning notes. As in the HE control arm, there were no significant differences among SOAP sections within or across the embedding models.

#### 3.3.2. Fused Sensor and SOAP Embeddings Features

As shown in the *Fall AE* column group in Table **7(a)**, fused BioBERT features resulted in perfect AUROC for Fall AE when fit on Subjective notes (AUROC=1.0(1.0), Recall=1.0) and Objective notes (AUROC=1.0(1.0), Recall=1.0). Assessment (AUROC=0.92(0.90–0.95), Recall=0.74) and Planning (AUROC=0.98(0.97–0.99), Recall=0.85) performed well but with lower recall rates. The AUROC confidence intervals for Subjective, Objective, and Planning sections indicate that those features performed significantly better compared to Assessment features. ClinicalBERT fused features also had perfect AUROC for Fall AE on Subjective (AUROC=1.0(1.0), Recall=1.0) and Objective (AUROC=1.0(1.0), Recall=1.0) sections. Assessment (AUROC=0.92(0.90–0.95), Recall=0.74) and Planning (AUROC=0.97(0.96–0.99), Recall=0.85) were slightly lower. As with BioBERT, all models for other notes performed significantly better than Assessment. Fused BERT-Uncased features had similar perfect AUROC on Subjective (AUROC=1.0(1.0), Recall=1.0) and Objective (AUROC=1.0(1.0), Recall=1.0) sections. Assessment (AUROC=0.93(0.90–0.95), Recall=0.74) and Planning (AUROC=0.96(0.93–0.98), Recall=0.55) had lower discrimination and recall. The Subjective and Objective sections significantly outperformed both the Assessment and Planning models. Across all fused feature sets in the HE control arm, Fall AE models fit on Subjective and Objective notes produced perfect AUROC and recall, which was significantly better than Assessment sections for every model (BioBERT, BioClinicalBERT, BERT-Uncased). Planning notes also performed well, but did have marginal overlapping confidence intervals with Subjective and Objective sections for BioBERT and BioClinicalBERT models.

The intervention group models performed similar to the control cohort, reported in the *Fall AE* column group in Table **7b**, with fused BioBERT features having perfect AUROC when fit on Subjective notes (AUROC=1.0(1.0), Recall=1.0) and Objective notes (AUROC=1.0(0.99–1.0), Recall=0.99). Assessment (AUROC=0.99(0.99–1.0), Recall=0.93) and Planning (AUROC=1.0(0.99–1.0), Recall=0.95) also performed at or near perfect, with no significant differences among SOAP sections. Clinical-BERT fused features had similar results, with Subjective (AUROC=1.0(1.0), Recall=1.0), Objective (AUROC=1.0(0.99–1.0), Recall=0.99), Assessment (AUROC=0.99(0.99–1.0), Recall=0.93), and Planning (AUROC=1.0(0.99–1.0), Recall=0.96) notes having near perfect performance, again with no significant differences. The fused BERT-Uncased models resulted in Subjective (AUROC=1.0(1.0), Recall=1.0), Objective (AUROC=1.0(0.99–1.0), Recall=0.99), Assessment (AUROC=0.99(0.99–1.0), Recall=0.93) and Planning (AUROC=1.0(0.99–1.0), Recall=0.96) notes also showing similar near-perfect performance. Confidence intervals for AUROC overlapped across all SOAP sections. When comparing across feature sets, all models in the SM intervention arm had perfect or near-perfect discrimination for AE Occurrence regardless of the embedding or note type, and the overlapping confidence intervals indicate no significant differences among models or feature sets. However, Assessment and Planning notes demonstrated decreased recall across all models.

## 4. Discussion

The prediction of adverse events from clinical SOAP notes and remote sensor data features can potentially improve care coordination and guide patient interventions, requiring fewer clinical visits and assessments. In addition to negatively impacting participant health, adverse events can potentially disrupt a clinical trial study through reduced trial sampling. For this reason, the detection of adverse events is an important step to mitigating their effects on study safety. Of the adverse event types documented in this randomized clinical trial, those related to fall events were most common across both cohorts.

### 4.1. Comparing Documentation Practices in AE and Non-AE Notes

Results showed that AE notes were consistently shorter and contained fewer NER-mapped tokens than non-AE notes in both cohorts. Particularly in the Objective and Subjective sections, documentation text related to AEs was more concise, possibly reflecting an emphasis on reporting clinical aspects of the AE. The SM intervention cohort had higher documentation rates with more notes and higher token counts in nearly all sections compared to the HE controls, particularly in the Subjective section where the AE related notes had a higher token count than HE notes, despite being shorter than non-AE notes within the same cohort. This suggests that the presence of the intervention may have encouraged clinicians or patients to provide more detailed narrative accounts and patient-focused content, particularly regarding patient experience and perspective. The higher volume and detail in the SM notes together with the observed narrowing of AUROC confidence intervals in the model evaluation indicate that the intervention not only improved the quantity of information captured but also likely resulted in improved quality or standardized event documentation.

### 4.2. Performance Contrast Between Language Embedding Models

There were minimal difference between BioBERT, BioClinicalBERT, and BERT-Uncased models for both AE Occurrence and Fall Injury classification tasks. For models trained solely on text embeddings, all three models had comparable AUROC and recall across SOAP note sections, with overlapping confidence intervals indicating no statistically significant differences. Although BioBERT and Bio-ClinicalBERT, pretrained on biomedical or clinical corpus, were expected to better capture patterns from the SOAP notes, they performed comparable to the general-domain BERT-Uncased model. The BERT-Uncased had the highest AUROC compared to the BioBERT embeddings for both AE occurrence and fall events in the HE controls, and for fall events in the SM arm against both BioBERT and BioClinicalBERT embeddings, using Subjective notes. These results suggest that general language representations were sufficient to capture clinically relevant information from SOAP notes. The lack of a performance from domain-specific pretraining may reflect the presence of narrative patient-perspectives with non-medical language used in the SOAP notes. As such, model selection among these three embedding types could be based on factors such as computational efficiency or availability, rather than domain-specific pretraining alone. Future work could explore further improvements by fine-tuning a base BERT model directly on clinical SOAP notes for use in downstream tasks and language models with pretrained on clinical documentation or UMLS-specific corpora.

### 4.3. Performance Contrast Between Fused Feature Sets

For all three BERT-based models (BioBERT, BioClinicalBERT, and BERT-Uncased) non-fused models had more variable AUROC and recall, dependent on the SOAP note section. Subjective notes had the best overall performance among the non-fused models, but AUROC values didn’t approach near-perfect, and recall was inconsistent across models and arms. Additionally, AUROC confidence intervals overlapped between different note sections and BERT models, indicating that improvements were not always statistically better. However, integrating sensor features with BERT embeddings provided significant improvements in both discrimination and recall. Across both study arms, models fit on fused features had perfect or near-perfect AUROC and recall, particularly for Subjective and Objective note sections. The confidence intervals for these fused models were narrow and, in several cases, did not overlap even for the lower performing Assessment and Planning sections with the non-fused embeddings only counterpart models. Additionally, improvements were observed across all three BERT models evaluated, indicating that sensor integration enhances performance regardless of the specific language embedding model used. This contrast in performance between fused and non-fused feature sets illustrates the complementary relationship between the high frequency biophysical sensor data and clinical documentation texts. While BERT models extract clinical context from narrative notes, the sensor data contributes objective, continuous signals which when merged, enhanced event detection. The findings suggest that implementing models for AE detection should prioritize multimodal approaches over relying on narrative or biophysical sensor measures on their own.

### 4.4. Performance Contrast Between SOAP Components

Classifier performances across SOAP note sections were dependent on the event type (AE occurrence, fall) and SOAP section features. For AE occurrence models, Subjective and to a lesser degree, Objective, notes provided higher discrimination and recall across all BERT models tested. These sections provided better signals for identifying AEs likely due to the inclusion of patient-reported symptoms and direct clinical observations that precede or coincide with event onset. Assessment and Planning notes, in contrast, had lower and less consistent performance. This difference was more apparent for models fit on fused features, with Subjective and Objective sections having perfect or near-perfect AUROC and recall, while Assessment and Planning had lower performances, particularly in the HE control arm. For fall AE models, there was less difference in performance between SOAP components among the BERT models. None of the SOAP sections outperformed others across the embedding models for either study arm, and overall discrimination was lower compared AE occurrence classifiers. This may reflect that falls were more acute and documented less frequently in the SOAP notes, reflected by the sensor-only models outperforming the embedding models for fall AE detection. However, for models fit on the fused feature sets in both classification tasks, the Subjective and Objective sections had perfect or near-perfect AUROC and recall, while Assessment and Planning features were less predictive, though only marginally lower in the SM intervention cohort. The results indicate that Subjective and Objective notes were more informative for identifying AEs and fall events when fused with sensor data. Assessment and Planning sections, which contain more clinical reasoning and care strategies, contributed less to predicting either event. To improve detection of both general AE and fall event-specific tasks, future work may focus on extracting and engineering patient-summary features, possibly through fusion across varied language models or decision level, from Subjective and Objective documentation.

### 4.5. Performance Contrast Between Study Arms

The differences in model performances between the control (HE) and intervention (SM) study arms suggest that the intervention may have influenced the detectability of AEs, perhaps through the quality of clinician documentation as described previously. In non-fused models, Subjective notes for both arms were more predictive for AE occurrence, but the intervention arm had slightly higher AUROCs and, increased recall in some cases, as with BioBERT embeddings. Confidence intervals in the intervention arm were narrower and had wider differences in performance between note sections, particularly when compared against Subjective notes, indicating better signal and potentially improved clinical note quality related to the intervention. Fall AE models did not perform well with embeddings-only features, with sensor-based models consistently outperforming all of the embedding models. Performance in both arms dramatically improved by fusing embeddings with sensor features regardless of SOAP context. The SM intervention arm had near-perfect performance across all note types and embedding models, with overlapping confidence intervals indicating less difference between SOAP sections. However, for HE controls the differences between Subjective/Objective and Assessment/Planning sections after fusion were significant, having non-overlapping confidence intervals, suggesting that perhaps the documentation and event reporting were less uniform for the control group. Overall, the results indicate that the predictive signals were more consistent in the intervention arm, possibly due to increased clinical engagement or documentation standards due to the intervention, lending to improved event capturing. The SM cohort not only had higher performance across most models but also demonstrated more uniform results from all note types on fused features. These findings indicate that increased fidelity of clinical documentation combined with multimodal data integration meaningfully enhances the performance and generalizability of AE detection models.

## 5. Conclusion

This study evaluated the performance of classification models for predicting adverse event (AE) occurrence and fall injuries from clinical SOAP note embeddings (BioBERT, BioClinicalBERT, and BERT-Uncased) and in-home sensor data collected from rural older adult clinical trial participants. Across both control (HE) and intervention (SM) cohorts, there were only marginal differences in performance between non-fused embedding models, with BERT-Uncased performing as well or better than pretrained domain-specific models using Subjective note sections. These results suggest that general-domain language models are sufficient at extracting information related to AEs from clinical trial documentation, where biomedical pretraining may not capture relevant terms from more narrative or patient-perspective text as found in SOAP notes. Sensor-derived health features were less predictive of AE occurrence, but were better predictors for classifying fall injury events compared to the non-fused text embeddings. Fusing clinical text embeddings and in-home sensor features improved performance for both classification tasks, demonstrating that multimodal features enhance the predictive modeling of clinical event outcomes.

Additionally, the Subjective and Objective components of SOAP notes were found to be the most predictive for both classification tasks, suggesting that captured patient perspectives and direct clinical observations are important indicators for monitoring intervention progress or tracking changes in health status. Comparisons between the HE control and SM intervention cohorts showed that the intervention arm had higher documentation rates, longer and more detailed SOAP notes, and narrower AUROC confidence intervals, possibly reflecting more consistent and higher quality documentation. In contrast, the control cohort’s documentation was less detailed, and model performance was more variable across note sections. These differences indicate that clinician engagement and intervention documentation practices can directly improve the use of patient records for event modeling and prediction. These findings support the integration of narrative and physiological data streams for anticipating AEs and tailoring interventions, which potentially could reduce the frequency of in-person assessments for older adults. Future research may focus on model refinement using text embedding models more specific to NER extraction and UMLS corpus matching and exploration of additional multimodal fusion techniques. These efforts could further advance the use of remote monitoring and predictive analytics to other patient populations, such as those experiencing functional decline, providing support for proactive care across clinical settings.

## Ethics Statement

This work involved human subjects or animals in its research. Approval of all ethical and experimental procedures and protocols was granted by University of Missouri Institutional Review Board under Application No. 2043542.

## Author Contributions

NM: Conceptualization, Data Curation, Formal analysis, Investigation, Methodology, Software, Validation, Visualization, Writing – original draft, Writing – review & editing. MP: Conceptualization, Methodology, Supervision, Writing – review & editing. ER: Conceptualization, Funding acquisition, Project administration, Writing – review & editing. RW: Conceptualization, Funding acquisition, Project administration, Supervision, Writing – review & editing. SX: Conceptualization, Investigation, Methodology, Resources, Supervision, Writing – review & editing.

## Data Availability Statement

The data supporting this study are derived from human subjects and contain sensitive information; therefore, they are not publicly available. Access to de-identified data may be granted upon reasonable request to [Rachel Wolpert, Principal Investigator], subject to institutional review board approval.

## Acknowledgments

The University of Missouri has a financial relationship with Foresite Healthcare, who produces the sensor platform used in this study.

